# Ageing Biomarkers Derived From Retinal Imaging: A Scoping Review

**DOI:** 10.1101/2024.02.13.24302673

**Authors:** Michaela Grimbly, Sheri-Michelle Koopowitz, Alice Ruiye Chen, Zihan Sun, Paul Foster, Mingguang He, Dan J. Stein, Jonathan Ipser, Lisa Zhuoting Zhu

## Abstract

**Background/Aims:** The emerging concept of retinal age, a biomarker derived from retinal images, holds promise in estimating biological age. The retinal age gap (RAG) represents the difference between retinal age and chronological age which serves as an indicator of deviations from normal ageing. This scoping review aims to collate studies on retinal age to determine its potential clinical utility and to identify knowledge gaps for future research.

**Methods:** Using the PRISMA checklist, eligible non-review, human studies were identified, selected, and appraised. Pubmed, Scopus, SciELO, PsycINFO, Google Scholar, Cochrane, CINAHL, Africa Wide EBSCO, MedRxiv, and BioRxiv databases were searched to identify literature pertaining to retinal age, the RAG, and their associations. No restrictions were imposed on publication date.

**Results:** Thirteen articles published between 2022 and 2023 were analysed, revealing four models capable of determining biological age from retinal images. Three models, ‘Retinal Age’, ‘EyeAge’ and a ‘convolutional network-based model,’ achieved comparable mean absolute errors (MAE): 3.55, 3.30 and 3.97 respectively. A fourth model, ‘RetiAGE’, predicting the probability of being older than 65 years, also demonstrated strong predictive ability with respect to clinical outcomes. In the models identified, a higher predicted RAG demonstrated an association with negative occurrences, notably mortality and cardiovascular health outcomes.

**Conclusion:** This review highlights the potential clinical application of retinal age and RAG, emphasising the need for further research to establish their generalisability for clinical use, particularly in neuropsychiatry. The identified models showcase promising accuracy in estimating biological age, suggesting its viability for evaluating health status.

**Key Message:** *What is already known on this topic:* Retinal age has emerged as a promising ageing biomarker capable of determining biological age from retinal images.

*What this study adds:* This study presents a comprehensive scoping review of current literature concerning retinal age and the RAG, highlighting the reproducible association between advanced retinal age gap and increased mortality and cardiovascular disease risk.

*How this study might affect research, practice, or policy:* The findings underscore the paucity of knowledge on this topic, advocating for further research in this area to determine the potential clinical use of retinal age as a biomarker.

**Synopsis:** This scoping review explores retinal age, a biological ageing marker derived from retinal images. Findings from 13 articles reveal links between advanced retinal ageing gaps and adverse outcomes, emphasizing its potential for improving health outcomes.

## Introduction

Globally, the number of individuals aged over 60 is rising, leading to an increased burden to healthcare services and society. Ageing changes are heterogenous, with substantial variation in the health impacts of ageing across populations, individuals and tissues (Lowsky et al., 2014; Tian et al., 2023). Thus, biological ageing markers have emerged to better represent the ageing process and predict functional capability.

Retinal age, an imaging based biomarker, provides an estimate of biological age derived from retinal fundus photographs (Nusinovici et al., 2022; Zhu, Shi, et al., 2023). The rationale for relying on retinal age as a biomarker stems from the retina’s shared embryological origin with the central nervous system (Ptito et al., 2021) and microvascular structure, which is closely related to that of the brain, heart and kidney (Elias et al., 2016; Farrah et al., 2020; London et al., 2013). Although retinal imaging has largely been used by ophthalmologists for improved understanding and treatment of ocular disease (Keane & Sadda, 2014), predictive retinal ageing extends this use by applying deep learning to retinal fundus photography. This greatly enhances the utility of retinal imaging beyond the realm of Ophthalmology.

The introduction of the retinal age gap (RAG), defined as the difference between calculated retinal age and chronological age, provides a valuable metric for assessing deviations from normal ageing. When compared to traditional biomarker approaches, often criticised for their cost, invasiveness, time-consuming nature, and suboptimal accuracy, the application of retinal age models provides a cost-effective, non-invasive, and readily accessible way of estimating biological age (Butler et al., 2004; Chen, Wang, et al., 2023). This makes it particularly suitable for large scale population studies.

To date, there is no specific review on the reliability of retinal age as a biomarker. Although several biomarker reviews have included retinal age as one of many biological age estimators, these reviews have not been able to provide a comprehensive summary of the accuracy, practical utilisation, or relevance of retinal ageing models in the healthcare domain. This scoping review seeks to consolidate what is known about retinal age, while identifying gaps for future research by collating studies published on retinal age.

Specifically, this review aims to answer the following questions:

- How extensive is the current literature pertaining to retinal age and RAG?
- How many models exist, and how accurate are they?
- What are the clinical associations of retinal age and RAG?
- Does this biomarker exhibit clinical utility?
- What are the most pressing future directions for research?

## Methods

### Protocol and registration

A scoping review is well suited to expand and consolidate what is known about the retinal age, as it allows synthesis of current literature. A protocol was developed for this purpose and registered on Open Science Forum (Available at: https://osf.io/fse75/ DOI 10.17605/OSF.IO/FSE75). The format for this scoping review is based on the PRISMA-ScR checklist (Page et al., 2021) and has made use of the Janna Briggs Institute Manual for Evidence Synthesis (Peters et al., 2017).

### Eligibility criteria

To ensure a broad search of current literature, all published literature, and preprints of primary studies of the retinal age in adults were included for analysis. There were no limitations imposed for publication language or publication date.

### Search

A comprehensive literature search of the following electronic databases was conducted from 17 June 2023 – 19 June 2023: Pubmed, Scopus, Cochrane Library, CINAHL, SciELO, Google Scholar, PsycINFO, Africa Wide EBSCO Host, MedRxiv and BioRxiv. A librarian assisted with formulating the search strategy. Initial search terms for the Pubmed database included, “retinal age” AND “association”, which were further refined to ((retinal age [Text Word]) OR (retinal age gap [Text Word])) AND (((association) OR (link)) OR (biomarker)) and adapted for each database searched. Please refer to appendix 1 for full search strategy used.

### Selection

A two-stage selection process was employed to assess relevance of studies identified in the search. Articles identified through the above-mentioned search strategy were deduplicated, after which two reviewers (MG, SK) independently screened titles and abstracts to ascertain eligibility and relevance, utilising the predetermined inclusion and exclusion criteria described above. For publications that met inclusion criteria, these criteria were re-applied to the full-text articles. In cases where discrepancies in reviewers’ ratings were observed, a coordinating investigator (JI) conducted a final review to determine inclusion eligibility. The citations within included articles were also scanned to identify any additional articles that may be suitable for inclusion. Email updates for Google Scholar were enabled to capture newly published literature between the initial search date and the writing phase. Subsequent articles found were subjected to the same review process.

### Data charting process

Data was extracted from the articles deemed suitable for inclusion using a spreadsheet developed by the reviewers. Data extracted included title, publication date, authors, study design, aim of the study, type of retinal age model used, training and validation details of the model, outcome of interest, analytical approach used, and key findings of the papers.

## Results

### Selection of sources

The PRISMA flow diagram (Figure 1) outlines the article selection process. The search strategy yielded a total of 342 articles (Appendix 1). After initial application of inclusion criteria, a total of 41 full text articles were examined for eligibility. Additionally, four articles published after the literature search date were considered for inclusion. A total of 13 articles met criteria for inclusion in the scoping review.

**Figure 1.**
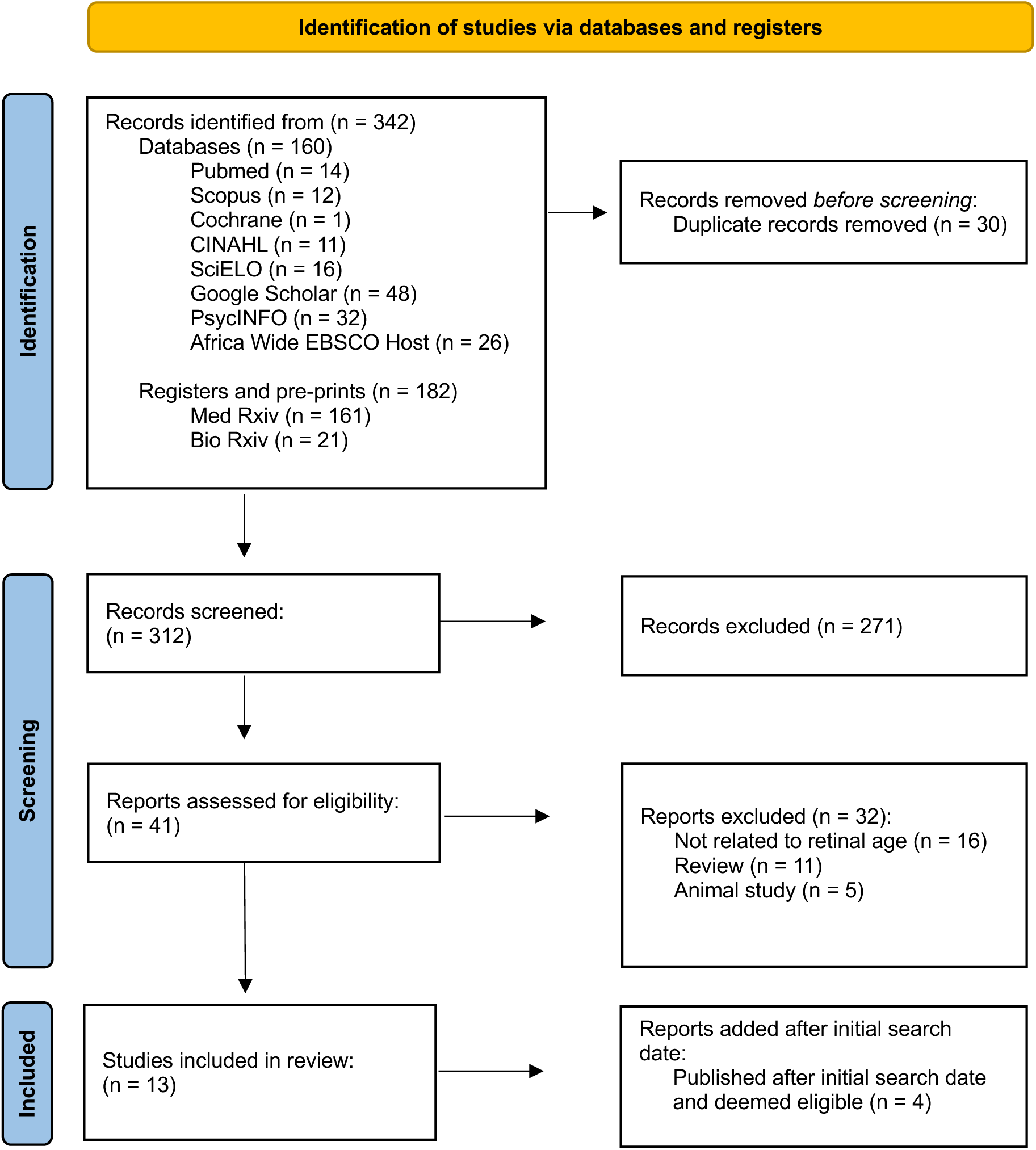
PRISMA 2020 flow diagram

### Characteristics of the Articles

All articles included for review were published between 2022 and 2023. Appendix 2 presents a summary of the studies.

### Narrative Review of Study Findings

All included studies utilised deep learning algorithms for retinal age analysis based on retinal fundus photographs. Four distinct models capable of determining biological age from retinal fundus photographs are outlined, with their training, validating, and testing processes described below. This is followed by a description of the application of these models to clinical populations.

#### Model Development and Accuracy

Three models were designed to predict age from retinal images, namely: ‘**Retinal Age’** (Zhu, Shi, et al., 2023), ‘**EyeAge’** (Ahadi et al., 2023) and a ‘**convolutional network-based model’** (Abreu-Gonzalez et al., 2023). A fourth biological ageing model, ‘**RetiAGE’** (Nusinovici et al., 2022), was developed to estimate the probability of an individual being older than 65 years from a retinal image.

The **‘Retinal Age’** model was trained and validated on 19 200 fundus photographs from 11 052 healthy UK Biobank participants. The model underwent 5-fold cross validation and achieved a mean absolute error (MAE) of 3.55, and Pearson correlation coefficient (R) between estimated age and chronological age of 0.80 (Zhu, Shi, et al., 2023). The **‘EyeAge’** model was trained on 217 289 images from 100 692 individuals in the EyePACS dataset, validated on 54 292 images from 25 238 individuals the same dataset and tested in both the UK Biobank and EyePACS dataset. The model achieved a MAE of 3.30, and a Pearson R of 0.87 for the UK Biobank test dataset, with corresponding figures of 2.86 and 0.95 for the EyePACS dataset (Ahadi et al., 2023). The **‘convolutional network-based model’** was trained on 98 400 photos from patients diagnosed with diabetes who were enrolled in the Retisalud programme of the Canary Islands Health Service. One thousand images from the dataset were used to validate this model, achieving a MAE of 3.97. However, it is unclear whether these validation images were included in the initial training set. The **‘RetiAGE’** model was trained on 116 312 photographs from 36 432 participants of the Korean Health Screening Study and validated on 12 924 unseen photos from 4 048 participants from the same dataset. In an internal test on 32 318 photos of 10 171 participants, the model achieved an Area Under the Receiver Operating Characteristic (AUROC) curve of 0.968 (95% confidence interval (CI): 0.965-0.970) and an area under the precision-recall curve (AUPRC) of 0.83 (95% CI: 0.83-0.84). When applied to the UK Biobank cohort, the model achieved an AUROC of 0.756 (95% CI: 0.753-0.759) and an AUPRC of 0.399 (95% CI: 0.388-0.410) with a correlation of 0.62 between ‘RetiAGE’ and chronological age (Nusinovici et al., 2022).

#### Clinical Utility and Model Associations

The RAG has conventionally been used as the metric for assessing the clinical utility of retinal age. Defined as the difference between the retinal age biomarker and chronological age, the RAG is used to assess the performance of the retinal age model in reflecting ageing. Eleven papers utilising two models, ‘**Retinal Age’** and **‘convolutional network-based model’**, have been published with the RAG as the outcome measure of interest.

Ten association analyses were conducted using the **‘Retinal Age’** model to explore the relationship between RAG and age-related parameters, within the UK Biobank cohort. The introductory study, highlighting the development of the model, revealed a significant association of a 2% increase in mortality risk for each one-year increase in RAG [Hazards Risk (HR) = 1.02, 95% CI 1.00-1.03, p=0.020] (Zhu, Shi, et al., 2023). Beyond the risk stratification for mortality, several prospective studies have highlighted associations for each one-year increase in RAG with a 10% increase of Parkinson’s disease [HR=1.10, 95% CI: 1.01-1.20, p=0.023] (Hu et al., 2022), a 4% increase of stroke [HR=1.04, 95% CI: 1.00-1.08, p=0.029] (Zhu, Hu, et al., 2022), a 3% increase of incident cardiovascular disease [HR = 1.03, 95% CI: 1.00-1.05, p=0.019] (Zhu, Chen, et al., 2022), a 10% increase in risk of incident kidney failure [HR = 1.10, 95% CI: 1.03-1.17, p=0.003] (Zhang et al., 2022), and a 7% increased risk of diabetic retinopathy in diabetic patients [HR = 1.07, 95% CI: 1.02-1.12, p=0.004] (Chen, Zhang, Hu, et al., 2023). Several cross-sectional studies utilising the **‘Retinal Age’** model explored the associations between certain lifestyle diseases and RAG. Central obesity [p<0.001] (Chen, Zhang, Shang, et al., 2023), higher glycemia levels [p<0.001] (Chen, Xu, Zhang, et al., 2023) and metabolic syndrome and inflammation [Odds Ratio = 1.01, 95% CI: 1.00-1.02, p= 0.0016] (Zhu, Liu, et al., 2023) were associated with higher RAGs, while a study aimed at correlating the RAG with cardiovascular health metrics, composed of 7 metrics totalling a score out of 14, determined that each 1-unit score increase in cardiovascular health was associated with a 13% decrease in calculated RAG [OR = 0.87, 95% CI: 0.85-0.90, p<0.001] (Chen, Xu, Shang, et al., 2023).

The **‘EyeAge’** model evaluated its clinical performance by calculating EyeAge Acceleration, determined akin to RAG, as the difference between EyeAge (reflecting retinal age) and chronological age. The acceleration is derived through regressing EyeAge against chronological age for assessment of the model’s clinical performance. In the UK Biobank cohort, adjusted EyeAge achieved an all-cause mortality hazard ratio of 1.03, while RAG (referred to as EyeAge Acceleration) was associated with a higher risk of chronic obstructive pulmonary disease [p=0.0048], myocardial infarction [p=0.049], elevated systolic blood pressure [p=1.025e-7] and fluid intelligence scores [p=5.34e-27]. Additionally, a genome wide association study (GWAS) was performed as part of the study and found that candidate genes identified for EyeAge acceleration are associated with eye function and age-related disorders (Ahadi et al., 2023).

The Abreu ‘**convolutional network-based model**’ determined, from a case-control study, that in diabetic patients from the Retisalud programme of the Canary Islands, a higher RAG was associated with more severe, progressive diabetic retinopathy [p<0.001] (Abreu-Gonzalez et al., 2023), echoing the findings of the **‘Retinal Age’** model in diabetic patients (Chen, Zhang, Hu, et al., 2023).

The **‘RetiAGE**’ model also used the UK Biobank cohort to assess its clinical performance. In contrast to ‘**Retinal Age’** and the ‘**convolutional network-based model,**’ which utilised calculated RAG for outcome assessment, the developers of the ‘RetiAGE’ biological age marker directly investigated its association with different age-related outcomes. Individuals placed in higher quartiles of **‘RetiAGE’** were considered to have accelerated ageing. Comparing those within the fourth quartile group to those in the first quartile, there was a 67% higher risk of 10 year all-cause mortality [HR = 1.67, 95% CI: 1.42-1.95], a 142% higher risk of cardiovascular related mortality [HR = 2.42, 95% CI: 1.69-3.48] and a 60% higher risk of cancer related mortality [HR=1.60, 95% CI: 1.31-1.96] after adjustment for chronological age and other established ageing biomarkers (Nusinovici et al., 2022).

#### Saliency Maps

Features that drive the retinal ageing estimates were identified for the ‘**Retinal Age**’ and **‘RetiAGE’** models. **‘Retinal Age’** retrieved attention maps using guided Grad-CAM (Selvaraju et al., 2020), to highlight pixels in the input retinal fundus image based on their contributions to the final evaluation. Areas around retinal vessels are highlighted, indicating that retinal microvasculature is used by the deep learning model for age prediction (Zhu, Shi, et al., 2023). ‘**RetiAGE**’ generated saliency maps to localise anatomy contributing to retinal aging. They indicated that the model focuses on the macula, optic disc, and, echoing the findings from the **‘Retinal Age’** model, retinal vessels for age determination (Nusinovici et al., 2022).

## Discussion

This study aimed to qualitatively appraise existing research utilising retinal photography for the development of biological ageing markers. We sought to determine the accuracy of retinal age prediction models and evaluate their performance in reflecting age-related parameters, as well as their clinical associations. This scoping review identified several models which are able to estimate chronological age from retinal images with moderate to high accuracy, and identified several age-related associations. Here we present key findings from our review of the existing literature.

Four models are currently available to estimate biological age from retinal images, all based on deep learning algorithms: ‘**Retinal Age’** (Zhu, Shi, et al., 2023), ‘**EyeAge’** (Ahadi et al., 2023), the **‘convolutional network-based model’** (Abreu-Gonzalez et al., 2023) and ‘**RetiAGE’** (Nusinovici et al., 2022). The ‘**Retinal Age’**, **‘EyeAge’** and the **‘convolutional network-based model**’ were trained to predict numerical chronological age from retinal images, while ‘**RetiAGE’** was trained to predict the probability of an individual being over the age of 65 years.

All models were exclusively trained and validated using a single dataset, predominantly comprising Caucasian or Asian populations. To enhance robustness, both ‘**EyeAge’** and ‘**RetiAGE’** underwent additional internal testing on previously unseen images from the training and validation cohort. For model testing and outcome assessment, the UK Biobank cohort was utilised by three models: ‘**Retinal Age’**, ‘**EyeAge’** and ‘**RetiAGE.’** Although each of the four identified models demonstrated comparable model accuracy and performance, it is important to note that performance metrics were inconsistently reported, with some pertaining to validation performance, and others test performance. Consequently, the generalisability of these models is uncertain, with further work warranted to assess the applicability of these models across diverse populations.

Nevertheless, the application of retinal age models in predicting mortality and morbidity carries significant clinical implications. A key finding from these thirteen selected papers emphasizes that accelerated ageing, whether calculated as a RAG, age acceleration, or other indices, consistently correlates with mortality risk across three models (Ahadi et al., 2023; Nusinovici et al., 2022; Zhu, Shi, et al., 2023). In addition, ‘**Retinal Age’** and ‘**EyeAge’** show consistent associations with cardiovascular disease, while ‘**Retinal Age’** and the **‘convolutional network-based model’** show connections with the risk of diabetic retinopathy in diabetic patients. These findings highlight the potential of retinal age as an informative tool for quantifying the risk of mortality and cardiovascular morbidity. However, to date, no clinical trials have explored the utility or feasibility of the models, which is a crucial aspect for determining their clinical relevance. Furthermore, factors associated with higher RAG, including glycaemic status (Chen, Xu, Zhang, et al., 2023), central obesity (Chen, Zhang, Shang, et al., 2023) and metabolic syndrome (Zhu, Liu, et al., 2023), suggest that RAG may provide valuable insight into lifestyle habits and traits that accelerate ageing. Notably, these factors were only investigated using the ‘**Retinal Age’** model.

It is important to highlight the inadequate reporting of characteristics of the populations used for training the retinal age prediction models. Only the ‘**Retinal Age’** model mentions that it was trained on healthy populations. This distinction is important if one wishes to consider biological age as equal to chronological age in normal ageing individuals. The health status of the population used for training ‘**EyeAge’** and ‘**RetiAGE’** remains undisclosed, while the **‘convolutional network-based model’** used data from diabetic patients. This may lead to confounding the effects of diabetes on apparent aging with age itself. Such a discrepancy could lead to controversy over whether these three retinal age models are accurate predictors of biological age. Thus, there is a need for a standardized procedure of developing biological age which should be proposed for use in further studies.

Additionally, these models were trained on a limited set of retinal features with only two models, ‘**Retinal Age’** and ‘**RetiAge**’, producing saliency or attention maps to identify the potential features used by the deep learning models for age assessment. This links to concerns about regulatory compliance and interpretability of the use of artificial intelligence in health care (Albahri et al., 2023; Funer, 2022). However, both models alluded to retinal microvasculature being a key component of age ascertainment, indicating that retinal age may reflect ageing related to vascular status. This is supported by the finding that retinal age models are particularly associated with cardiovascular health (Chen, Xu, Shang, et al., 2023; Nusinovici et al., 2022). To improve understanding of retinal features that align with biological age, advanced or diverse visualization techniques are imperative.

The application of retinal age models in predicting neuropsychiatric diseases is relatively underexplored. Given that the retina is an extension of the central nervous system, it offers a unique and accessible ‘window’ to visualise cerebral neuronal health (Elias et al., 2016; Farrah et al., 2020; London et al., 2013). An extensive body of work has found that changes in the retina, most notably thinning in the retinal nerve fibre layer, may be associated with certain neuropsychiatric and neurodegenerative diseases (Gonzalez-Diaz et al., 2022; Kashani et al., 2021). In our review, only one paper using the ‘**Retinal Age’** model explored the RAG in the realm of neuropsychiatry, specifically in the context of Parkinson’s disease, leaving this area underexplored (Hu et al., 2022). As neurodegeneration is an important aspect of ageing, future studies should concentrate on improving our understanding of the connections between retinal age and neuropsychiatric conditions.

Several limitations of this scoping review deserve emphasis. Publications in non-indexed journals and other ‘grey literature’ may have been missed. Due to insufficient data availability, it was not possible to quantitatively synthesis data using meta-analytic statistical techniques. As more literature becomes available on the topic, conducting a broader, more extensive review may unveil more diverse associations of retinal age, mechanisms for associations, and possibly link retinal age to other biomarkers. Strengths of our study included its development according to a predefined protocol, and application of the PRISMA extension for scoping reviews.

In conclusion, this scoping review identified four retinal ageing models derived from retinal images, with advancing retinal age gaps significantly associated with mortality and cardiovascular disease. It highlights the scarcity of data in the realm of neuropsychiatry, emphasises the need for a standardised procedure in developing retinal ageing models, and shows that testing across different datasets is crucial to improve the generalisability and utility of the models. Improving our understanding of the biological underpinnings of how these models determine age may too improve their reliability in reflecting ageing processes. Nevertheless, the evidence highlights the potential of retinal age as a biomarker, suggesting its viability as a valuable, cost-effective tool for evaluating health status.

## Data Availability

All data produced in the present work are contained in the manuscript

## Funding

This research received no external funding.

## Supplementary Materials

## Appendix 1

### Search of Published Works Databases

Preliminary search strategy formulated on Pubmed.

Initial search terms included: retinal age, retinal age gap, association, link, biomarker.

| Database | Date of Search |  | Search Terms | Results | Articles Screened |
| --- | --- | --- | --- | --- | --- |
| Pubmed | 17/06/2023 | 1 | (retinal age) OR (retinal age gap) | 49 442 | 14 |
|  |  | 2 | (retinal age[Text Word] OR (retinal age gap[Text Word])) | 23 |  |
|  |  | 3 | (retinal age[Text Word] OR (retinal age gap[Text Word]) AND (((association) OR (link) OR (biomarker))) | 14 |  |
| Scopus | 17/06/2023 | 1 | Retinal AND age OR retinal AND age AND gap | 9 430 | 12 |
|  |  | 2 | TITLE-ABS-KEY ("retinal age" OR "retinal age gap") | 29 |  |
|  |  | 3 | TITLE-ABS-KEY (association OR biomarker OR link)) AND TITLE-ABS-KEY ("retinal age" OR "retinal age gap") | 12 |  |
| Cochrane | 17/06/2023 | 1 | (retinal age OR retinal age gap):ti,ab,kw (Word variations have been searched) | 5362 | 1 |
|  |  | 2 | (retinal age OR retinal age gap):ti,ab,kw AND (association OR link OR biomarker):ti,ab,kw (Word variations have been searched) | 1402 |  |
|  |  | 3 | ("retinal age" OR "retinal age gap" OR "retina age"):ti,ab,kw AND (association OR link OR biomarker):ti,ab,kw NOT (age related macular degeneration):ti,ab,kw (Word variations have been searched) | 1 |  |
| CINAHL | 17/06/2023 | 1 | "retinal age" OR "retinal age gap" | 3 | 11 |
|  |  | 2 | "retinal age" AND "association" | 11 |  |
| SciELO | 17/06/2023 | 1 | Retinal age | 223 | 16 |
|  |  | 2 | Retinal age gap | 0 |  |
|  |  | 3 | Retinal age OR retinal age gap | 0 |  |
|  |  | 4 | Retinal age AND association | 16 |  |
| Google Scholar | 18/06/2023 | 1 | "retinal age" OR "retinal age gap" AND association OR link OR biomarker | 48 | 48 |
| **PsychINFO** | 18/06/2023 | 1 | (retinal age OR retinal age gap) AND (association OR link OR biomarker)<br>Boolean/Phrase | 32 | 32 |
| **Africa Wide**<br>**EBSCO Host** | 18/06/2023 | 1 | (retinal age OR retinal age gap) AND (association OR link OR biomarker)<br>Boolean/Phrase | 26 | 26 |
| **Total identified from published databases** |  |  |  |  | 160 |

### Search of Preprint Databases

| Database | Date of Search |  | Search Terms | Results | Articles Screened |
| --- | --- | --- | --- | --- | --- |
| **MedRxiv** | 19/06/2023 | 1 | "retinal age gap" | 161 | 161 |
| **NIH Reporter** | 19/06/2023 | 1 | "retinal age gap" OR "retinal age" (advanced)<br>Limit to: Project Title, Project Terms, Project Abstract | 1 project<br>4<br>publications | 0 |
| **BioArchives** | 19/06/2023 | 1 | "retinal age" | 21 | 21 |
| **Total identified from preprint databases** |  |  |  |  | 182 |

## Appendix 2

**Table 1:**
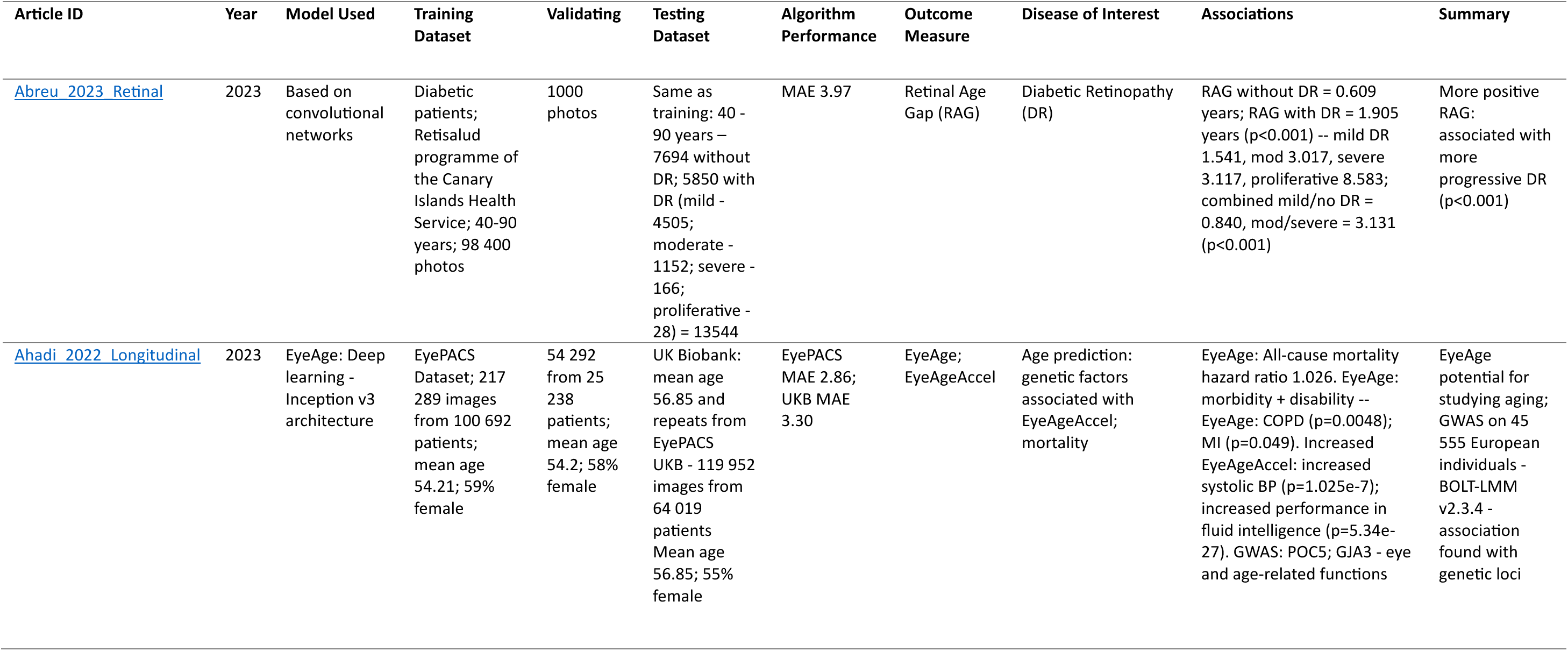
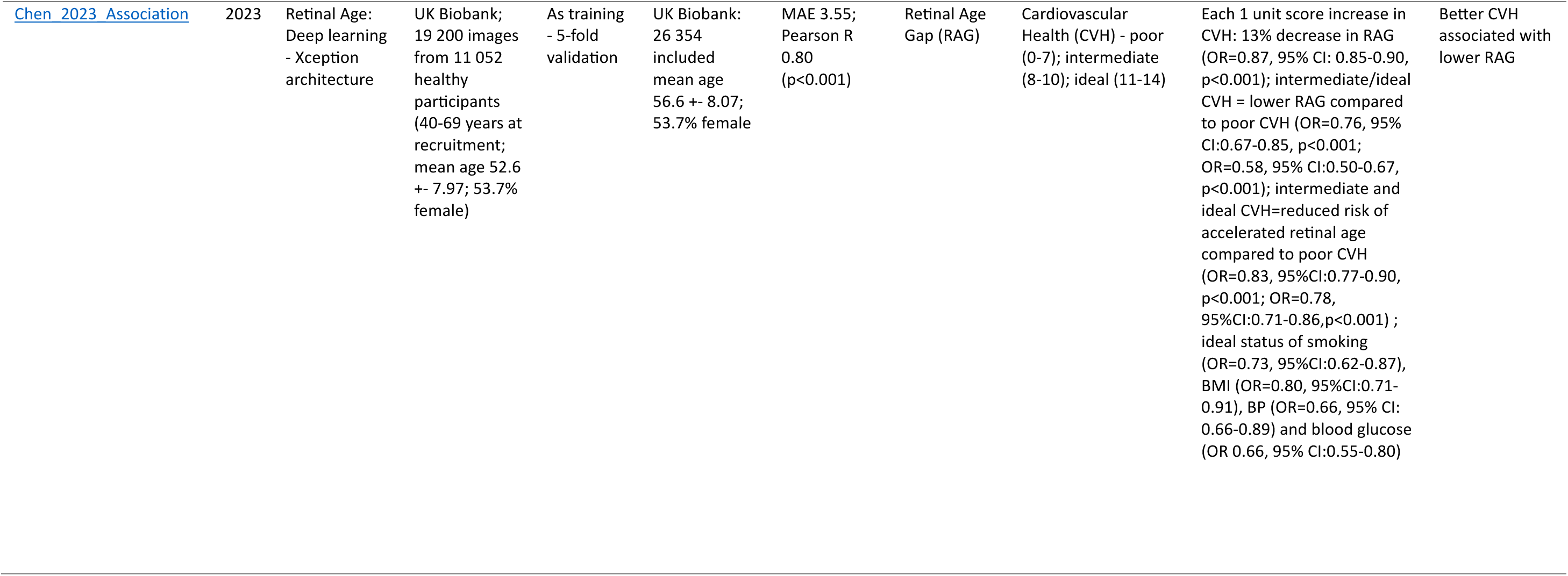
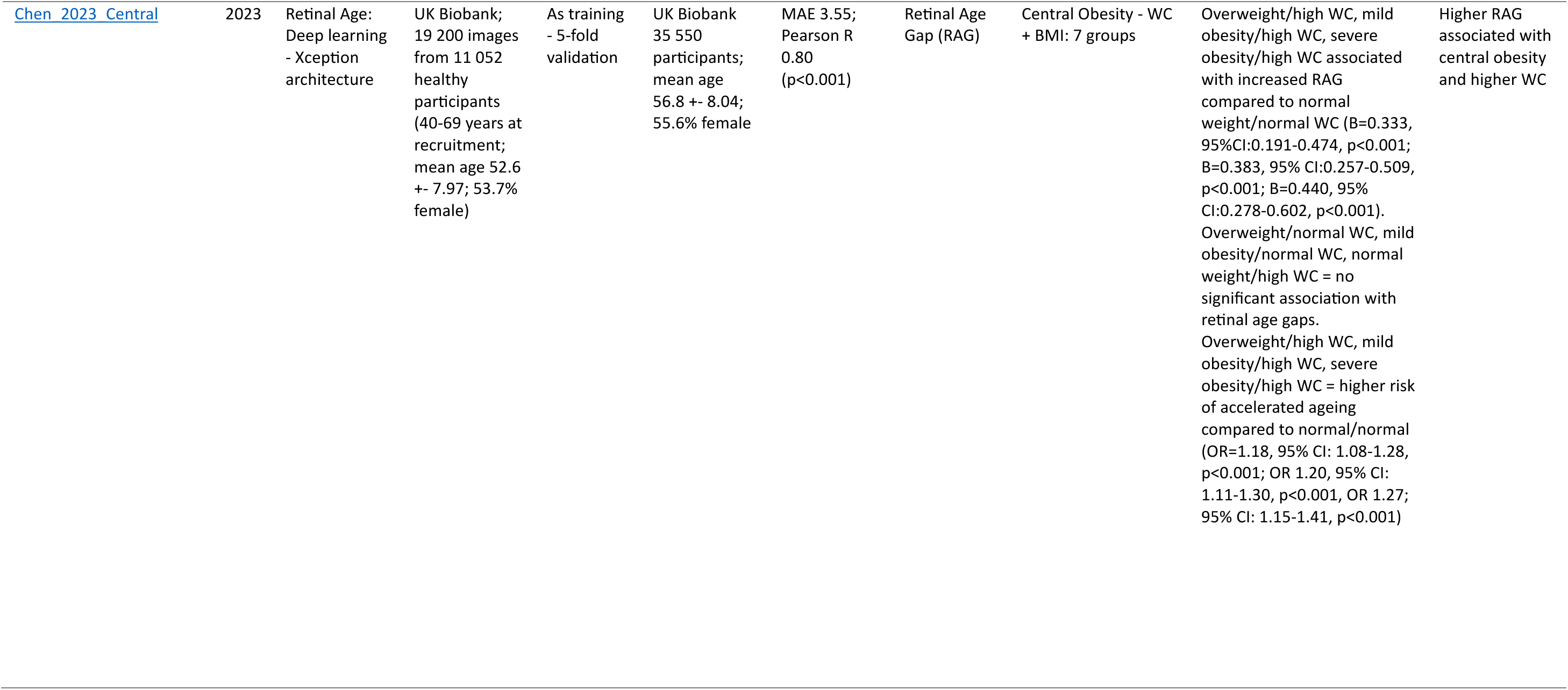
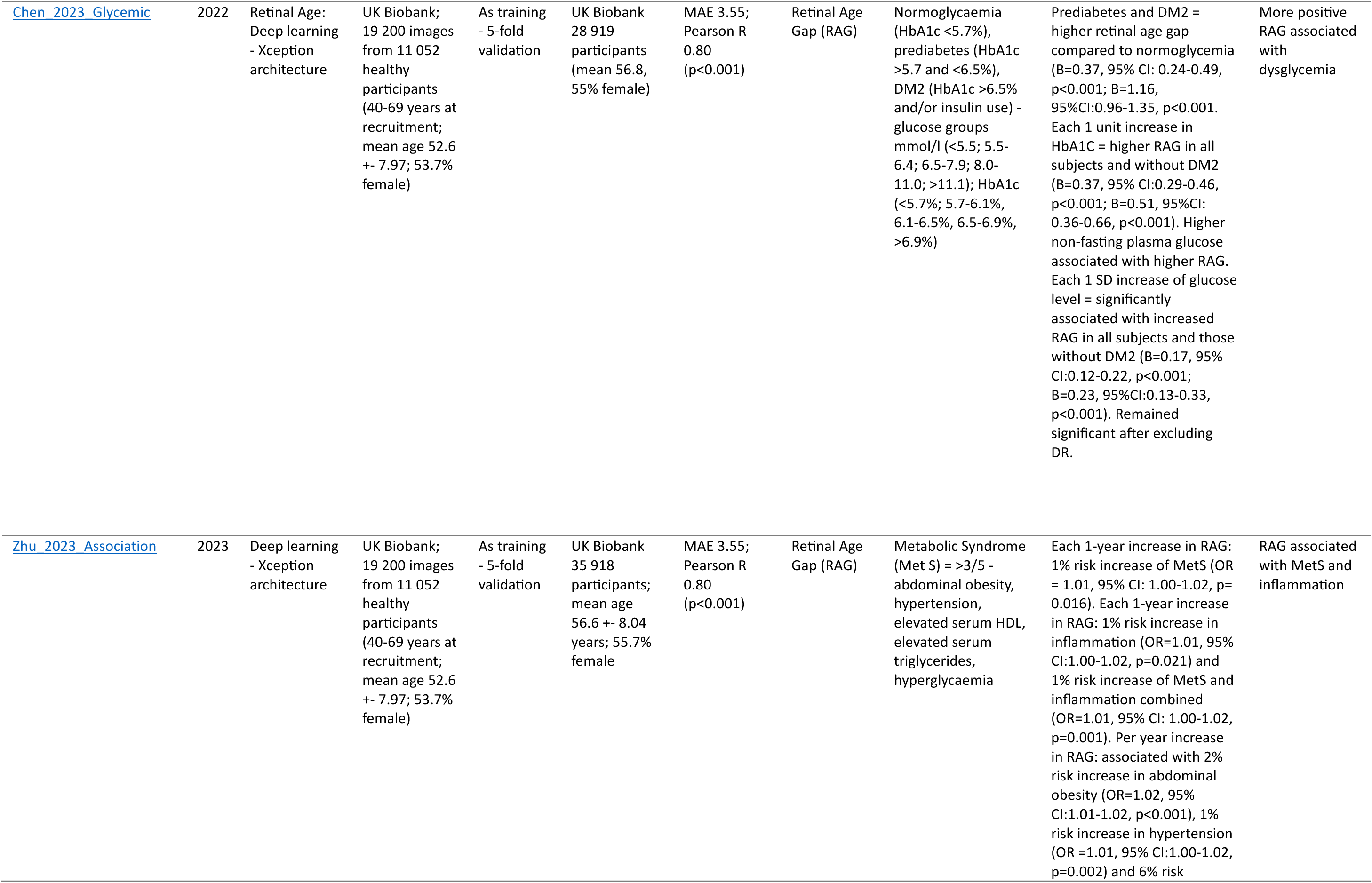
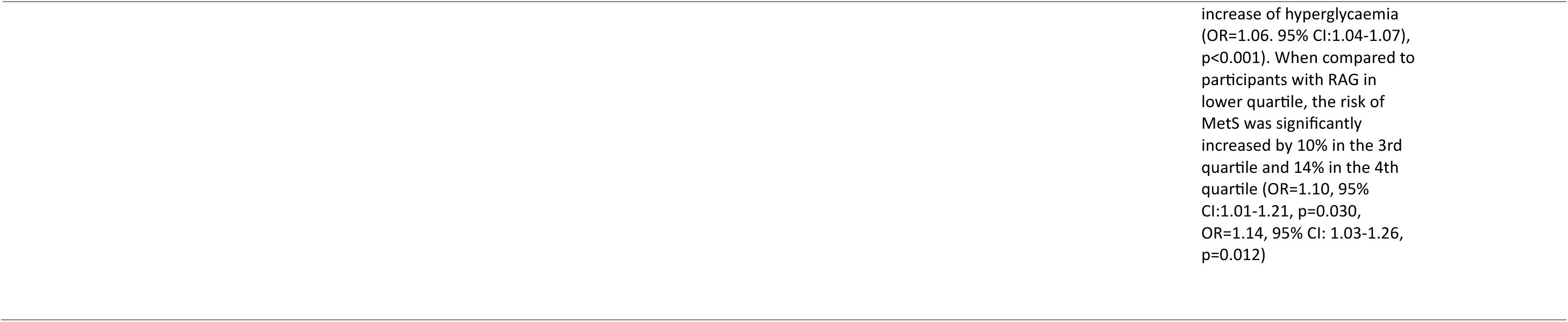
Cross Sectional Studies – Data Extraction.

**Table 2:** Prospective Studies – Data Extraction.

| Article ID | Year | Model Used | Training Dataset | Validating | Testing Dataset | Algorithm Performance | Outcome Measure | Disease of Interest | Associations | Summary |
| --- | --- | --- | --- | --- | --- | --- | --- | --- | --- | --- |
| Chen 2023 Retinal | 2023 | Retinal Age: Deep learning – Xception Architecture | UK Biobank; 19 200 images from 11 052 healthy participants (40-69 years at recruitment; mean age 52.6 +- 7.97; 53.7% female) | As training - 5-fold validation | UK Biobank 2311 diabetic patients without diabetic retinopathy (DR) at baseline. Mean age 58.5; 39.7% female | MAE 3.55; Pearson R 0.80 (p<0.001) | Retinal Age Gap (RAG) | Incident diabetic retinopathy | Each 1-year increase in RAG: 7% adjusted increase in risk of incident DR [HR=1.07, 95% CI: 1.02-1.12, p=0.004]. RAG in fourth quartile had higher DR risk [HR=2.88, 95% CI: 1.61-5.15, p<0.001]. DR risk increased across RAG quartiles [HR=1.35, 95% CI 1.11-1.64, p=0.002] | More positive RAG associated with increased risk of incident DR |
| Hu 2022 Retinal | 2022 | Retinal Age: Deep learning - Xception architecture | UK Biobank; 19 200 images from 11 052 healthy participants (40-69 years at recruitment; mean age 52.6 +- 7.97; 53.7% female) | As training - 5-fold validation | UK Biobank 35 834 participants free of Parkinson's disease (PD) at baseline; 56+-8.04; 55.7% female | MAE 3.55; Pearson R 0.80 (p<0.001) | Retinal Age Gap (RAG) | Incident Parkinson's disease - ICD 9/10 codes - history of PD; incident PD | Each 1-year increase in RAG: 10% increase in risk of PD (HR=1.10, 95% CI: 1.01-1.20, p=0.023). Compared with lowest RAG quartile, the risk of PD was increased in third and fourth quartiles (HR = 2.66, 95% CI: 1.13-6.22, p=0.024; HR = 4.86, 95% CI: 1.59-14.8, p=0.005). Predictive value of retinal age model (AUC = 0.708, 95% CI:0.638-0.778) and risk factor model (AUC=0.717, 95% CI:0.633-0.802) was similar (p=0.821). | More positive RAG associated with future risk of incident PD |
| Nusinovici 2022 Retinal | 2022 | RetiAge: Visual Geometry Group (VGG) - deep convolutional neural network architecture | Korean Health Screening Study; 129 236 photos from 40 480 participants; mean age 54 +- 9.01 | 12 924 photos from 4 048 participants | 56 301 from UK Biobank (mean age 57.1 +- 8.3 years; 46.5% female) used for all-cause mortality, CVD mortality and cancer mortality.<br>46 551 from Korean Health Screening Study (mean 53.6 +- 9.2 years; 45.4% female) used for all-cause mortality. | AUROC 0.968 (95% CI: 0.965-0.970); AUPRC 0.83 (95% CI: 0.83-0.84) in internal test<br>AUROC of 0.756 (95% CI: 0.753-0.759) and an AUPRC of 0.399 (95% CI: 0.388-0.410) in UK Biobank test | RetiAge | Predict old age from retina. All-cause mortality; CVD mortality and cancer mortality | Independent of chronological age and phenotypic ageing markers: when compared to RetiAge first quartile, those in the RetiAge fourth quartile had 67% higher risk of 10 year all-cause mortality (HR = 1.67, 95% CI: 1.42-1.95, p <0.001) 142% higher risk of CVD mortality (HR = 2.42, 95% CI: 1.69-3.48, p<0.001) and 60% higher risk of cancer mortality (HR=1.60, 95% CI: 1.31-1.96, p<0.001). | RetiAge can predict aging; higher RetiAge associated with mortality risk |
| Zhang 2023 Association | 2023 | Deep learning - Xception architecture | UK Biobank; 19 200 images from 11 052 healthy participants (40-69 years at recruitment; mean age 52.6 +- 7.97; 53.7% female) | As training - 5-fold validation | UK Biobank 35 864 participants with no kidney failure at baseline; mean age 56.75 +- 8.04; 55.7% female | MAE 3.55; Pearson R 0.80 (p<0.001) | Retinal Age Gap (RAG) | Incident kidney failure - ICD 10 and OPCS 4 codes | Each 1-year increase in RAG: 9% increase in risk of incident kidney failure (HR = 1.09, 95% CI: 1.03-1.15, p<0.001). Retinal age gaps in the highest quartile had a significantly higher risk of incident kidney failure compared to those in the first quartile (HR = 2.77, 95% CI: 1.29-5.93, p=0.009). There was a graded association of incident kidney failure across retinal age gap quartiles (p=0.004). Participants in higher RAG quartile showed higher risk of kidney failure event. | More positive RAG associated with incident kidney failure |
| Zhu 2022 Association | 2022 | Deep learning - Xception architecture | UK Biobank; 19 200 images from 11 052 healthy participants (40-69 years at recruitment; mean age 52.6 +- 7.97; 53.7% female) | As training - 5-fold validation | UK Biobank 35 541 participants; mean age 56.8 +- 8.04; 55.6% female | MAE 3.55; Pearson R 0.80 (p<0.001) | Retinal Age Gap (RAG) | ASI - arterial stiffness index; CVD events - cardiovascular disease | Each 1-year increase in RAG: increase in ASI (B=0.002, 95% CI: 0.0001-0.003, p=0.001). Higher odd of having severe ASI with a larger RAG (OR=1.01, 95% CI: 1.01-1.02, p<0.001). Each 1-year increase in RAG: predicts 3% increase in risk of incident CVD (HR = 1.03, 95% CI: 1.00-1.05, p=0.019). Remains significant when ASI is incorporated (HR = 1.03, 95% CI: 1.01-1.06, p=0.019) | More positive RAG significantly associated with ASI. RAG predictor of future risk of incident CVD. |
| Zhu 2022 Retinal | 2022 | Deep learning - Xception architecture | UK Biobank; 19 200 images from 11 052 healthy participants (40-69 years at recruitment; mean age 52.6 +- 7.97; 53.7% female) | As training - 5-fold validation | UK Biobank 35 304 without stroke history; mean age 56.7 +- 8.04; 55.9% female) | MAE 3.55; Pearson R 0.80 (p<0.001) | Retinal Age Gap (RAG) | Stroke - ICD 9/10 codes | Each 1-year increase in RAG: associated 4% increase in risk of stroke when adjusting for confounding factors (HR=1.04, 95% CI: 1.00-1.08, p=0.029). Retinal age gaps in the 5th quintile had higher risk of stroke compared to those whose RAG was in the first quintile (HR=2.37, 95% CI:1.37-4.10, p=0.002). Predictive capability of retinal age alone in 10-year stroke risk was comparable to a well-established risk factor-based model (AUC=0.676 vs AUC 0.661, p=0.511) | More positive RAG associated with incident stroke |
| Zhu 2023 Retinal | 2023 | Deep learning - Xception architecture | UK Biobank; 19 200 images from 11 052 healthy participants (40-69 years at recruitment; mean age 52.6 +- 7.97; 53.7% female) | As training - 5-fold validation | UK Biobank 35 913 photos from 35 917 participants; mean age 56.8 +- 8.04; 55.7% female | MAE 3.55; Pearson R 0.80 (p<0.001) | Retinal Age Gap (RAG) | All-cause mortality | Each 1-year increase in RAG: 2% increase in mortality risk (HR = 1.02, 95% CI 1.00-1.03, p=0.020); Positive retinal age gap: substantially increased mortality; Higher retinal age gap (3rd and 4th quartile): higher non-CVD/cancer associated death (HR=1.49, 95% CI 1.13-1.96, p=0.005; HR=1.67, 95% CI 1.17-2.39, p=0.005) | More positive RAG associated with increased risk of mortality (>non-CVD/non-cancer) |

## Notes

### Competing Interest Statement

The authors have declared no competing interest.

### Clinical Protocols

https://osf.io/fse75/

### Funding Statement

This study did not receive any funding

